# What medical students think about measurement of their wellbeing and the implications for pastoral support: cross sectional survey and qualitative interviews

**DOI:** 10.1101/2021.08.24.21262526

**Authors:** G Simons, R Effah, DS Baldwin

## Abstract

**Objectives:** To find out how, why and when medical students think wellbeing should be measured.

**Design:** A mixed methods study comprising a cross-sectional online survey (November 2020-March 2021) and semi-structured on-line interviews. Views on the frequency of availability for measurement, the format, type and purpose of measurement, and with whom wellbeing should be discussed were measured. When an outcome was scored 7-9 on a 9-point Likert scale of agreement by ≥75% of participants it was considered critical, in line with COMET and GRADE processes for rating recommendations. Inductive thematic analysis was undertaken on the interview transcripts by two independent researchers.

**Setting:** All Medicine programmes at University of Southampton.

**Participants:** Medical students from all years took part in the survey (n=118) and interviews (n=16).

**Results:** Participant demographics were similar to national medical student demographics. Most participants (94%) felt able to give 5 minutes to measure their wellbeing at least once a month. No single format of measurement was rated critical. Research, governance and individual feedback all reached the 75% threshold for the purpose being considered critically important. Only subjective assessments undertaken by the individual in real-time were rated as critically important (78.1%) measurement tools. Students selected that they would discuss their wellbeing with other medical students (n=87) nearly as often as they selected a member of the Faculty (n=104). Top determinants of wellbeing picked by medical students were energy, ability to do activities of daily living, and negative feelings. Five interview themes further explained these findings.

**Conclusions:** Five recommendations about self-care teaching, quality-assured pastoral and peer support, proactive wellbeing check-ins and demographic data are discussed in light of these findings. Methods to achieve them are suggested, which are medical student-centred, and which make use of existing resources.

**Strengths and limitations of this study:**

- This study provides new information on how, why and when medical students think their wellbeing should be measured.
- A mixed methods approach allowed the reasons behind the survey answers to be captured in ensuing interviews.
- Using the ≥75% a prior cut off for critical importance from COMET and GRADE allowed evidence-based recommendations for wellbeing measurement for medical students.
- It was not possible to recruit the number of students needed to make national inferences, although participant demographics were similar to national medical student demographics.
- A national cross-sectional survey would provide further evidence for medical schools in devising wellbeing provisions.

## Introduction

Mental health problems are the most commonly declared diagnoses on medical student provisional registration applications to the General Medical Council (GMC) (1). This is not surprising as 75% of mental health diagnoses are established by the age of 24 years (2). Addressing wellbeing at Medical School could help reduce the significantly higher levels of depression and anxiety seen in doctors compared to the general UK population (3–7). The GMC has recognised this in its promoting wellbeing guidance in “Supporting medical students with mental health conditions”(8). However, it does not recommend how educational strategies for wellbeing should be evaluated.

Medicine challenges student wellbeing more than most courses for several reasons. Recruitment policy inclusivity (9) has not yet caught up with content and support, with learning resources still being adapted to the widening demographics of medical students. The transition from college to university (10, 11) is harsher for often perfectionist medical students (12), as they move from being the highest-achieving big fish in a small pond to an average fish in a large shoal (13). ‘Imposter syndrome’ (14). The culture of competition, rather than collaboration, can detrimentally impact the wellbeing of medical students (15) and future team culture in the National Health Service (NHS). The length of the course leads to more students facing financial stressors, especially in the context of widening participation and graduate entry (16), and places further pressure on educational achievement. Medicine has the highest total workload hours of all degree courses (17), leaving less time for part-time work and wellbeing activities.

Medical students face a second transition from pre-clinical to clinical education (18). Keeping students’ clinical exposure at the desired level of difficulty (19) and not allowing it to tip into an unsafe experience for them, or patients, is challenging. The need to provide not only enabling services (20), but also occupational health (21), for example, is unique to healthcare students. Generic university pastoral support provisions are often not medicine assessment literate (22), nor equipped to cope with issues that might occur on placements with NHS partners. The role models who teach clinically are typically not formally trained educators (23), whereas formally trained University staff are often not doctors and may therefore lack insight into common clinical situations, such as exposure to traumatic events.

All students require a safe learning environment (19) that is open (24) but these most basic needs have been hard to meet at University in the context of a pandemic. Necessarily abrupt changes such as the move to online learning were shown to be anxiety provoking (25) for practical content such as anatomy, uncertainty about the loading of practical skills into periods when lockdown rules did not apply, and concerns whether online assessment would allow progression and graduation were unsettling (26). Compounding this, the loss of peer support on how to deal with uncertainty, moving from comprehension of uncertainty management, to application of it, is hard without social interaction (27). The pandemic has been harder for healthcare courses which had to send students into direct contact with the virus on placement (28).

In medical education, it is not yet standard practice for pastoral care to be evaluated and quality assured. The focus tends to be on pathology, when problems have already occurred, such as failed assessments, or mental health diagnoses, but this stifles evidence about which contexts allow students to thrive. There is no international consensus definition of wellbeing (29) and many different outcomes are measured to capture wellbeing with differing tools (30), making it hard to compare studies, or know how to evaluate educational interventions. To address this gap in knowledge and involve medical students in the development of medical education and evaluation tools (22), this study aims to establish, through medical student survey and interview, when, how, why, what, and by who, wellbeing should be measured.

## Methodology

A mixed methods study comprising a cross-sectional survey and subsequent semi-structured interviews.

### Cross sectional surveys

Reported using the STROBE guidelines (31).

### Sample

Medical students attending the University of Southampton (UOS) were recruited between November 2020 and March 2021. Students enrolled on any Medical degree programme in any year were eligible to participate.

### Data collection

Students were recruited using social media posts (Twitter, Instagram and Facebook), and “shout outs” prior to the start of lectures. Students were provided with a link to the survey hosted on the online survey platform “I-survey”.

### Outcomes

- Demographics (Year, programme, age, sex, ethnicity, religion)
- Frequency of availability to spend 5 minutes measuring wellbeing
- Format of wellbeing measurement
- Reasons for wellbeing measurement
- Type of wellbeing measurement
- Who to talk to about wellbeing
- Determinants of wellbeing

### Measurement

Office for National Statistics 2011 census questions were utilised for demographics. To assess agreement, 9-point Likert scales were used. No assumptions were made about student preferences, so free text options were available.

### Bias

Selection bias was mitigated as students were recruited through digital and non-digital routes.

### Study Size

The split of opinion amongst 37,500 medical students nationally (32) on the questions asked is unknown, so to account for anything between a 50/50 split to an 80/20 split, with a 95% confidence interval and ±5% sample error, between 245 and 381 surveys needed to be completed to allow national inferences (33).

### Quantitative variables

Where a 9-point Likert scale was used, and an outcome was scored 7-9 by 75% of participants it was considered critical. This was based on the use of ≥75%, as an acceptable cut off by a number of published studies looking to reach a consensus on outcome measurement (34–37), and accords with COMET (38, 39) and GRADE (40) processes for rating recommendations.

### Statistical methods

To account for where data was missing the *n* was reported for each question individually.

### Semi-structured interviews

Reported using SRQR recommendations (41).

### Qualitative approach

Constructivist epistemology, based on the concept that knowledge is built from experiences and social interactions was used in this project. Constructivism does not require knowledge to be deduced using one method, and several methods may be used to demonstrate something is “true”. Constructivism allows for more than measurable evidence, as required in Positivism, to represent external reality, and therefore allows the use of interviews (42).

### Researcher characteristics

A fourth year MMedSci student was trained to conduct the semi-structured interviews. As their relationship with participants was that of a peer, rather than senior, a greater level of trust was anticipated.

### Context

The on-line interviews took place via Microsoft Teams in line with Public Health England (PHE) guidance.

### Sampling strategy

Any participants that consented to being invited to interview in the online survey were approached. Sampling was stopped when thematic and meaning saturation were reached.

### Data collection methods

The interviewer followed a semi-structured interview schedule. The interviews were recorded on Microsoft Teams, transcribed using Microsoft Stream and cleaned with VTT Cleaner.

### Data processing

Interview recordings were stored in a limited access folder on the secure University network, available only to the research team. Transcripts were labelled with a participant number and any personal identifiers were removed.

### Data analysis

Inductive analysis was used to allow the medical students to generate the themes rather than impose an existing framework.

### Techniques to enhance trustworthiness

Themes were identified independently by two researchers (43) using “NVivo” qualitative analysis software. Triangulation of themes between reviewers, with the survey data and existing literature was undertaken (44, 45).

### Ethics

Ethical approval was obtained from the University of Southampton Research and Governance Office, Study number: 55730. All participants accessed a Participant Information Sheet and gave consent prior to taking part in the survey, and interviews. It was identified as a risk that students might be distressed by thinking about their wellbeing, and to mitigate this risk participants were given details of the BMA 24/7 confidential counselling and peer support service (0330 123 1245), the Samaritans (116 123), and advised to contact their GP if they were concerned about their mental health. Following the interview, a £10 Amazon voucher was emailed to each participant to compensate them for their time.

### Patient and Public Involvement

The research questions, study design and recruitment processes were designed with a medical student. PPI is at the heart of this study as its aim is to find out when, how, why, what, and by who, medical students think wellbeing should be measured. Dissemination of the results was undertaken collaboratively with a medical student.

## Results

### Surveys

A total of 118 medical students participated and were included, a 9.5% response rate from the total medical student population (n=1245). The demographics of the 118 participants are shown in Table 1.

**Table 1.** Demographics of the medical students in this survey compared to national comparators.

| *Demographics* | *This survey* | *National comparator* |
| --- | --- | --- |
| ***Gender*** |  |  |
| *n* | 118 | 70,370 <sup>1</sup> |
| % Male | 18.6 | 39.4 |
| % Female | 79.7 | 60.6 |
| %Prefer not to say | 1.7 | 0 |
| ***Age*** |  |  |
| *n* | 118 | 70,370 <sup>1</sup> |
| %18 – 20 | 24.6 | 31 |
| %21 – 24 | 63.6 | 37.6 |
| %25 - 29 | 11.8 | 15.4 |
| %30 and over | 0 | 16 |
| ***Ethnicity*** |  |  |
| *n* | 118 | 43605 <sup>2</sup> |
| %White/Other white background | 61.9 | 56.9 |
| % Mixed/Multiple ethnic groups | 5.9 | 5.2 |
| % Asian/Asian British Indian | 5.1 | 11.9 |
| % Asian/Asian British Pakistani | 3.4 | 7.7 |
| % Asian or Asian British Bangladeshi | 0.8 | 2.1 |
| % Chinese | 2.5 | 2.2 |
| % Other Asian background | 11.9 | 5.4 |
| % Black or Black British-African | 6.8 | 4.6 |
| % Black or Black British-Caribbean | 0 | 0.4 |
| % Other Black background | 0 | 0.1 |
| % Other ethnic groups | 0 | 3.4 |
| % Did not state | 1.7 | 0 |
| ***Religion*** | 118 | 2532385 <sup>2</sup> |
| % No religion | 39 | 49 |
| %Buddhist | 0.8 | 1 |
| %Christian | 30.5 | 32 |
| %Hindu | 7.6 | 3 |
| %Jewish | 0.8 | 0 |
| %Muslim | 10.2 | 10 |
| %Shinto | 0.8 | 0 |
| %Sikh | 1.7 | 1 |
| %Prefer not to say/not known | 8.5 | 4 |

<sup>1</sup> National data set from the Higher Education Statistics Agency, Medicine and Dentistry 2019/20 enrolment (46).
<sup>2</sup> National data set from the Higher Education Statistics Agency, all students enrolled 2019/20 (46).

When asked how often they could give 5 minutes to measure their wellbeing, 49.1% of answering participants (n=116) chose an option that was at least once a day, 78.4% an option that was at least once a week, and 94% an option that was at least once a month. This left 7 participants who could not give 5 minutes once a month.

No format of measurement was rated critically important (n=116). Surveys as downloaded apps, or online, were the only 2 formats with <15% rating them of limited importance. Some core outcome set studies use ≥15% ratings of limited importance as a cut-off to exclude options (47). Using this method face-to-face, phone or video call conversations, as well as paper surveys, would have been excluded as formats for wellbeing measurement.

When asked who they would feel comfortable discussing their wellbeing with at a 30 minute conversation, 42 participants of those that responded (n=95) selected that they would prefer to use a website or an app, with 6 participants saying they would not want to discuss wellbeing at all.

Students could select more than one option and selected an individual chosen by them (n=55), Personal Academic Tutors (n=50) and other medical students (n=87):more than generic university services (n=33), clinical supervisors (n=32) and the BMA (n=27).

Research, governance and individual feedback all reached the ≥75% threshold for the purpose of wellbeing measurement being considered critically important (Table 2). Only subjective measures taken by the individual in real-time, such as the 12-item General Health Questionnaire were rated critically important (78.1%) as a feasible, valid and reliable type of measure of wellbeing (Table 3).

**Table 2.** Medical student ratings for the purpose of wellbeing measurement in medical students. On the 9 point Likert scale the boundaries were categorised as follows: Limited Importance = 1-3, Important = 4-6, Critically important = 7-9.

| **Purpose** | **Limited Importance (%)** | **Important (%)** | **Critically important (%)** |
| --- | --- | --- | --- |
| **Research (n=101)** | 0 | 13.9 | 86.1 |
| **Governance nationally (n=101)** | 3 | 7.9 | 89.1 |
| **Governance locally (n=101)** | 3 | 3.9 | 93.1 |
| **Individual feedback (n=100)** | 0 | 16 | 84 |
| **Patient safety (n=101)** | 30.7 | 28.7 | 40.6 |
| **Introduction to exploring wellbeing (n=99)** | 1 | 27.3 | 72.7 |

**Table 3.** Medical student ratings of whether the types of measure of wellbeing might be feasible, valid and reliable in medical students. On the 9 point Likert scale the boundaries were categorised as follows: Limited Importance = 1-3, Important = 4-6, Critically important = 7-9.

| Type of measurement (n=96) | Limited Importance (%) | Important (%) | Critically Important (%) |
| --- | --- | --- | --- |
| A biomarker (e.g. hair cortisol levels) | 36.5 | 35.4 | 28.1 |
| A measure taken by someone else (e.g. sickness absence days) | 37.5 | 37.5 | 24 |
| A measure taken by you (e.g. Public health surveillance wellbeing scale) | 5.2 | 24 | 70.8 |
| A descriptive measure taken by you (e.g. reflective writing about your wellbeing over the last 12 months) | 16.8 | 30.6 | 52.6 |
| A measure taken by someone else in real-time (joined a Teams teaching session that day) | 25 | 36.5 | 38.5 |
| A measure taken by you in real-time *e.g. 12 item General Health Questionnaire GHQ12) | 5.2 | 16.7 | 78.1 |
| A descriptive measure taken by you in real-time (e.g. a daily blog) | 10.4 | 29.2 | 60.4 |

The top 4 determinants of wellbeing that should be measured which were chosen by participants (n=95) were: energy and fatigue (94.7%); the ability to do activities of daily living (92.6%); negative feelings (84.2%); and sleep and rest (81.1%).

### Interviews

55 participants gave consent to be contacted, and 16 interviews were undertaken. Themes and meanings identified:

#### 1. Wellbeing is mental wellbeing

Everyone thought of mental wellbeing, when asked to define wellbeing, with fewer thinking of physical, social or financial aspects, even in the context of a global pandemic.

> *“So, when people talk about wellbeing, I guess the first thing that I think about is mental wellbeing*.*” (Participant 2)*
>
> *“I mean we’re at uni and people love spouting on about mental health because it’s obviously a big issue. The mental health comes into mind for me, probably because I’ve always been healthy, like I’ve never had any serious illness*.*” (Participant 13)*

#### 2. Exercise and support from friends and family are most important for wellbeing

When asked about what positively impacted wellbeing the top two were exercise, particularly outside, and support from friends and family.

> *“You know, I’m a believer, like, the world is your kind of gym, so I like going to the Common when working out*.*” (Participant 5)*
>
> *“So, talking to my friends and my boyfriend helps with my wellbeing quite a lot and just like checking in with my family*.*” (Participant 9)*

#### 3. Isolation and the design of the Medicine Programme are detrimental to wellbeing

Students could not access their usual support networks during lockdown periods, including the 2020 Summer and Winter holidays if they were international students. Due to clinical placements, students were afraid of infecting others even where social interactions were allowed. Structural aspects of the medicine degree (such as exam timetabling, revision timetabling, who can ‘sign off’ clinical skills, competitive assessment, the length of the course) were all cited by participants as things that negatively impacted their wellbeing. Students reported having to stop the exercise they found so positive for their wellbeing for deadlines and exams.

> *“Normally, living alone is fine because I see my friends, but*.. *I didn’t want to have a support bubble because again, cross contamination. So, I spent the majority of my final year alone. In hospital, you know, you shouldn’t be like seeing friends*.
>
> *You shouldn’t be eating lunch together. So, I spent the majority of the year completely by myself*.*” (Participant 3)*
>
> *“I stopped exercising to help revise for finals” (Participant 1)*
>
> *“I guess, like, the course being so long makes you feel like you’ve invested so much of your life into this, that you just actually have to pass like*.*” (Participant 15)*

#### 4. There are advantages to surveys, and conversations to measure wellbeing

Surveys were perceived as quicker but less pressured, allowing reflection and flexibility around when they are undertaken. Conversations were valued for the empathy the other person might show, the opportunity for the other person to pick up on non-verbal cues and ask more.

> *“A survey, I can do it anytime and I could choose to do it like waiting for the bus… If they were talking to me, they would probably get more information out of me than if I was to do a survey” (Participant 1)*

#### 5. Personal academic tutors and medical students in later years are best to discuss wellbeing with

Participants valued the rapport established with Personal Academic Tutors (PAT) and where there was rapport, felt comfortable discussing their wellbeing with them. Some participants had to change PAT to achieve this. Participants valued the fact that students in later years would have recently experienced the same things, but were concerned that medical students might not be equipped to discuss wellbeing. Senior Tutors were viewed as unfamiliar and only able to signpost.

> *“Me personally, I’ve had the same personal academic tutor since year one. We get on really well. I, surprisingly, I’m very open about my wellbeing issues with him. I feel like there’s no like hierarchy between us. He’s been very, like, non-judgmental, and open and kind and generous with me…And, and, I feel comfortable speaking to him. (Participant 3)*
>
> *“Just someone on a similar level to me, or in a higher year and has gotten through it. I just think I’d value their advice because I’ve just got that knowledge that they’re going through a similar experience that I’m going through*.*” (Participant 8)*

## Discussion

### Self-care needs to be integrated into the curriculum and assessed

The GMC “Outcomes for graduates 2018” recommends that curriculums include how to “self-monitor, self-care and seek appropriate advice and support”(48). The fact that 17 students reported they could not give 5 minutes to record their wellbeing any more frequently than once a month and that the top determinants of wellbeing picked were basic needs according to Maslow’s hierarchy (49) would suggest self-care needs to be taught.

Exercise was the most common activity used to help mental wellbeing in a national study of medical students during the pandemic (50) and the same was found in interviews in this study. However, the students interviewed reported stopping exercise ahead of exams and deadlines. To meet World Health Organisation recommendations for physical activity and reduce symptoms of anxiety and depression and improve cognitive health and sleep (51), students need to be taught that they will have deadlines and pressures throughout their careers and how to prioritise self-care, to allow them to work into their 70s (52).

A connected programme design (53) that introduces and builds on Self-Care using a constructivist approach and spiral learning (54) could be employed. This model allows interleaving and spacing, which assists learners in differentiating new, difficult concepts (55) such as ‘moral injury’ (56).

Teaching based on cases created by students, would allow reverse mentoring, through staff being made aware of the new challenges which students face. For strategic learners like medical students, assessment would not only drive effort (22, 57, 58), but is diagnostic and dialogic allowing ‘correct as you go’ feedback (59) and dynamic tailoring of programme design in response to what students say in their assessments (11). This could prevent the negative impact of programme design on wellbeing reported in the interviews.

#### Suggested Self Care Assessments

**Reflective writing** at the end of each module, or at the time of critical events, covering learning outcomes on self-monitoring, self-care and support sought, where appropriate.

**Objective Structured Clinical Examinations (OSCEs)** that allow medical students to be the person running a Health Education England recommended wellbeing check-in (60), or attending one, with learning outcomes around communication skills, signposting and boundary setting, or self-monitoring and self-care respectively. These could be undertaken with other health and social care students.

### Pastoral support should be quality assured by students for students

Participants wanted to pick who they interacted with about their subjective real-time wellbeing (Table 3) and no single format to do this was critically important, suggesting the need for choice. Participants felt it was critically important for wellbeing to be measured for governance as well as research and individual feedback (Table 2). Pastoral support has not traditionally been subject to governance, but with the emphasis on quality assurance educationally (22) this seems like an oversight. This need not be work intensive or behaviourist (54), as the responsibility for establishing, measuring and upholding the expected standards for pastoral support could lie with students. Reflection on their self-care, interaction with PATs, and other services, and feedback on those interactions could form part of the previously suggested Self Care assessments. Collation and analysis of this feedback could be part of an andragogic process (61), an annual student research project (53). This would allow a process model in which the students have choice about how pastoral support is delivered (62) and an influence over content to ensure it is inclusive (63).

### Medical student peer support should be formalised and quality assured

Participants selected another medical student to discuss their wellbeing with more than PATs, generic University services, or national wellbeing services. Interviews revealed that isolation negatively impacted wellbeing, but students did have concerns about how equipped other students would be to deal with wellbeing discussions, raising the need for quality assured peer support, with clear boundaries. The GMC recommends that “*where medical schools want to put a formal peer support programme in place, they must make sure that those who provide the service are properly trained for and supported in this role*”(8).

### Demographics beyond gender and ethnicity must be captured

Demographic variables should be captured to understand the population that will be accessing the pastoral support. For example, being aware that 50% of participants had religious beliefs makes signposting students to placement partner chaplaincy services very relevant. Not only because they provide safe spaces for prayer and reflection for all, but also because they are very experienced in offering pastoral, spiritual and religious care after traumatic clinical events and are free (64).

### Limitations

The study lacked the power to enable national inferences, although the demographic characteristics of this sample are broadly similar to those of medical students nationally. In future studies disability data should also be captured, as nationally 11.8% of medical students declare a disability on entry (46) and this has implications for workforce and service planning. Widening participation data should also be captured in future work using the Higher Education Statistics Agency questions to plan adequate provision of enablement and financial services (46).

### Unique contribution and future research

Unlike other investigations of medical student wellbeing before (21), and during (65) the pandemic, this study made no assumptions about how wellbeing should be measured, allowing student preference to be captured. This study provides evidence to inform a Core Outcome Set for medical student wellbeing, an agreed minimum sets of outcomes that will allow research study results to be compared and synthesised (65). This study ensured what stakeholders value was captured (39, 66) a key part of Core Outcome Set development.

## Conclusions

Medical students thought that the measurement of their wellbeing was critically important for research, governance and individual feedback, showing their support for quality assurance of pastoral and peer support. They wanted to be able to choose surveys, or conversations, to measure their wellbeing, as well as the person they discussed their wellbeing with. The type of measurement viewed as critically important was subjective, experienced, quantitative questionnaires, supporting their comfort with frequent wellbeing measurement. The determinants of wellbeing rated the most important, and the insights from interviews, together highlight the need for Self-Care to be an integrated and assessed part of the medical curriculum. Solutions to deliver this have been recommended that are medical student-centred and make use of existing resources. This work may be transferable across health and social care degree programmes.

## Supporting information

Strobe Checklist

## Data Availability

All data supporting this study are openly available from the University of Southampton repository at (awaiting DOI) after thesis submission in January 2022, as the work forms part of a PhD.

## Acknowledgements

We thank Ms Aimee O’Neill and Professor Julia Sinclair for their contributions to discussions of notions of wellbeing and their connotations.

